# Associations between indicators of socioeconomic position and DNA methylation: A systematic review

**DOI:** 10.1101/2021.01.21.21250199

**Authors:** Janine K. Cerutti, Alexandre A. Lussier, Yiwen Zhu, Jiaxuan Liu, Erin C. Dunn

**Author notes:** **Corresponding Author:** Erin C. Dunn, ScD, MPH, Psychiatric and Neurodevelopmental Genetics Unit, Center for Genomic Medicine, Massachusetts General Hospital, 185 Cambridge Street, Simches Research Building 6th Floor, Boston, MA 02114.; website: www.thedunnlab.com.

## Abstract

Socioeconomic position (SEP) is a major determinant of health across the life course. Yet, little is known about the biological mechanisms explaining this relationship. One possible explanation is through an epigenetic process called DNA methylation (DNAm), wherein the socioeconomic environment causes no alteration in the DNA sequence but modifies gene activity, gene expression, and therefore long-term health. To understand the evidence supporting a potential SEP-DNAm link, we performed a systematic review of published empirical findings on the association between SEP (from prenatal development to adulthood) and DNAm measured across the life course, with an eye toward evaluating how the timing, duration, and type of SEP exposure influenced DNAm. Across the 37 studies we identified, there was some evidence for the effect of SEP timing and duration on DNAm, with early-life SEP and persistently low SEP being particularly strong indicators of DNAm. Different indicators of SEP also had some unique associations with DNAm profiles, suggesting that SEP is not a singular concept, but rather that different aspects of the socioeconomic environment can shift DNAm patterns through distinct pathways. These differences with respect to SEP timing, duration, and type were notable because they were detected even among heterogenous study designs. Overall, findings from this review underscore the importance of analyzing SEP timing, duration, and type, given the complex relationship between SEP and DNAm across the lifespan. To guide future research, we highlight current limitations in the literature and propose recommendations for overcoming some of these challenges.

## Introduction

Socioeconomic position (SEP) is commonly measured across health-related fields (1). It is a multidimensional concept, encompassing diverse social and economic components, such as actual resources (e.g., weekly income) and rank-based characteristics (e.g., occupational prestige) (2, 3). These components can be measured at the individual or aggregate level (e.g., household, neighborhood), and are often quantified by indicators such as education, occupation, and living conditions (1, 4, 5).

SEP is frequently studied because it is considered “a fundamental cause of disease” (6, p. 87). Decades of observational studies have shown that low SEP is strongly associated with adverse behavioral and health outcomes among children and adults, including the 14 major cause-of-death categories worldwide (6, 7, 8, 9, 10, 11, 12, 13). Evidence from experimental and quasi-experimental studies also suggests that SEP may play a *causal* role in driving these outcomes. Indeed, interventions and policies that provide food (14), housing (15), medical-care subsidies, (16) or income-transfer supplements (17) reflect widespread positive effects on health, emotional, behavioral, educational, and employment outcomes, as well as reductions in risks for psychiatric disorders, substance use, and criminal behavior. As one example, a natural experiment of children whose families received annual income supplements showed a 40% decrease in child psychopathology (18) compared to the four years before receiving supplements, with the protective effect of financial assistance persisting into early adulthood (19).

Although prior studies have established SEP as a potent determinant of health, the biological mechanisms explaining this relationship are not well understood. One widely pursued hypothesis is that SEP may change gene activity, gene expression, and subsequently long-term health (without altering DNA sequences) through the process of DNA methylation (DNAm), wherein methyl groups are added to cytosine bases in DNA, typically in cytosine-guanine (CpG) dinucleotides (20, 21). Recent human epigenetic studies suggest that SEP at the individual-, household-, or neighborhood-level may have lasting effects on the genome through DNAm alterations in both childhood (22, 23) and adulthood (24, 25). However, systematic reviews of this evidence base have been limited, due to their often narrow scope.

Here, we performed a systematic review of empirical studies investigating the association between SEP and DNAm in humans, focusing on multiple SEP indicators and DNAm measures across the life course. This comprehensive scope distinguishes our review from prior efforts, which explored a narrow subset of SEP exposure (e.g., neighborhood disadvantage) (26), mechanisms to investigate epigenetic changes (e.g., epigenetic clock, telomere attrition) (27), or specific time periods in the life course (e.g., early-life SEP) (28, 29, 30). Our overarching goal was to characterize the overall evidence on the relationship between SEP and DNAm to address two specific research questions, which have not yet been addressed in prior reviews.

First, does the timing and/or duration of SEP influence DNAm patterns? Prior studies have shown that low SEP is especially harmful when experienced early in development and chronically throughout childhood (12, 19, 31). However, we are unaware of any attempts to systematically search and compare findings between epigenetic studies that have investigated SEP timing and duration features. Answers to this research question will not only provide a better understanding of how aspects of the socioeconomic environment become biologically embedded across the lifespan, but will also help to guide future research imperatives to facilitate targeted intervention efforts aimed at reducing the negative sequelae of low SEP. Such insights could also help identify whether there are age stages or sensitive periods in development of elevated plasticity when SEP-induced DNAm changes are most likely to occur.

Our second research question was: do different SEP indicators show differential DNAm profiles? Studies analyzing multiple SEP indicators have found that individual SEP exposures may play related yet distinct roles in health and behavioral outcomes (32, 33, 34). However, to our knowledge, no studies have systematically examined whether there is converging evidence across studies that different SEP indicators associate with distinct patterns of DNAm changes. Discerning whether such patterns exist can help reveal whether different SEP exposures initiate similar (or distinct) biological pathways implicated in later disease outcomes.

## Methods

We performed this review in accordance with the Preferred Reporting Items for Systematic Reviews and Meta-Analyses (PRISMA) guidelines (35). Because the populations, research designs, and DNAm measures varied, we conducted a narrative synthesis, rather than a meta-analysis, of this literature to characterize the heterogeneity of studies and summarize patterns across studies (36, 37). We did not assess risk of publication bias, because most standard systematic review indices evaluating risk of bias are not applicable to the observational study types included here (38).

### Study Identification

We systematically identified articles published from inception through September 18, 2019 (date last searched) on PubMed and PsycINFO. We worked closely with an experienced reference librarian to develop a combination of database-specific index terms (e.g., ‘socioeconomic factors’, ‘epigenomics’) and individual terms located in the title or abstract (e.g., ‘income’, ‘occupation’, ‘epigenetics’), which were further refined through team discussion (see **Supplemental Materials** for final search terms). We also assessed reference lists of review articles and included additional relevant studies. The final results were exported into EndNote for evaluation.

### Study Selection

We included only human empirical studies that examined an independent association between SEP and DNAm, including global, candidate gene, and epigenome-wide approaches (see **Supplemental Materials** for inclusion and exclusion criteria). An independent reviewer evaluated the titles and abstracts of all publications identified by our search, and then reviewed the full texts of relevant publications to determine eligibility. We resolved any uncertainty on study eligibility by discussion with three other team members.

### Data Extraction Process

An independent reviewer extracted the data (in triplicate), discussed the results with team members, and continually updated the data in an iterative process based on team discussion. Three other independent reviewers verified data extraction results; any disagreements were resolved by consensus and team discussions. We extracted the following information from each study: 1) sample features (i.e., sample size, cohort name, sex, race/ethnicity, country of enrollment); 2) overarching research question and design; 3) SEP exposure features; 4) approach to analyzing DNAm (i.e., global, candidate gene, epigenome-wide association); 5) DNAm assessment age(s); 6) tissue type(s) investigated; 7) DNAm measurement method; 8) covariates; 9) SEP-DNAm associations examined; and 10) primary and secondary study findings. Of note, we defined “global” DNAm as measures or estimates of the overall DNA methylome, including methylation levels of repetitive elements in DNA (e.g., LINE-1 and Alu) (39).

To synthesize how studies conceptualized SEP, we categorized each SEP measure (referred to throughout this review as “indicator”) into one of the following domains: education, occupation, income, neighborhood, subsidy, composite (i.e., aggregated SEP measure), and other. Additionally, we reported how each SEP indicator was captured, specifically the method of data collection (e.g., subjective caregiver or self-report versus objective, taken from census-tract data or medical records, etc.; retrospective versus prospective) and also the measurement scale (e.g., dichotomous, categorical, continuous, or ordinal) used to classify individual low to high SEP status. Detailed information on SEP features is included in **Supplemental Materials**.

To more consistently compare results across studies, we extracted results of SEP-DNAm associations reported at the most stringent significance threshold within the simplest (or unadjusted) regression model. We recorded the direction of association with DNAm (lower SEP associated with increase/decrease in DNAm), if reported, in our main results. For epigenome-wide association studies (EWAS) that used the Illumina Human Methylation 450k array (450k array) method, we compiled all individual CpG site (CpGs) IDs analyzed and corresponding p-value and FDR/*q*-value. We contacted authors of seven studies for these summary statistics and retrieved the summary statistics online for the other two papers. No other contact with authors was made.

### Data Analysis

First, we conducted descriptive analyses to summarize characteristics of all studies included and reported average sample size, distribution of overall study characteristics, and individual-level study methods and results grouped by approach to analyzing DNAm. Next, we analyzed these studies in relation to our two research questions. To address our first question (Does the timing and/or duration of SEP influence DNAm profiles?), we performed a high-level comparison of DNAm findings between studies analyzing SEP exposure at more than one age. We addressed our second question (Do different SEP indicators show differential DNAm profiles?) in two parts using summary statistics from EWAS 450k array studies described above.

## Results

Our search returned 478 results; see **Figure S1** for flowchart of entire search and selection procedure. There has been a steady growth in studies over time investigating the relationship between SEP and DNAm (**Figure S2**). A total of 37 studies met our inclusion criteria, capturing global DNAm (n=7; **Table 1**) (40, 41, 42, 43, 44, 45, 46), candidate gene (n=18; **Table 2**) (24, 25, 47, 48, 49, 50, 51, 52, 53, 54, 55, 56, 57, 58, 59, 60, 61, 62), and EWAS (n=12; **Table 3**) (22, 23, 63, 64, 65, 66, 67, 68, 69, 70, 71, 72) studies. Detailed information on each study is provided in **Tables S1-S3**.

**Table 1.**
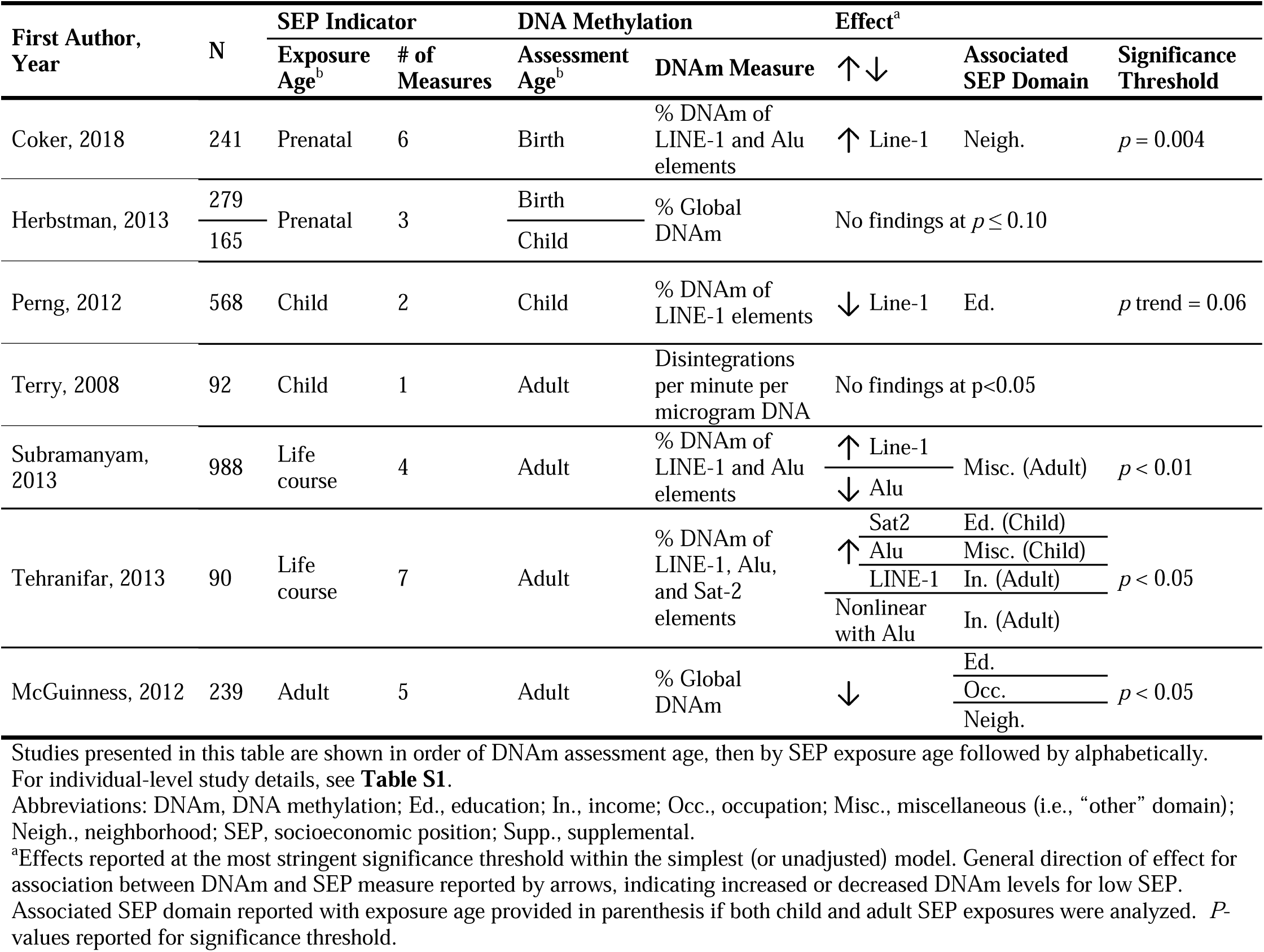
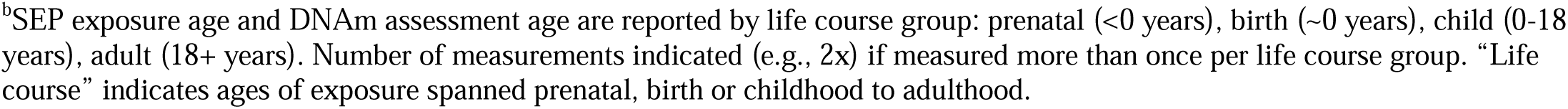
Associations Between Socioeconomic Position and DNA Methylation From Global DNA Methylation Studies (*n*=7)

**Table 2.** Associations Between Socioeconomic Position and DNA Methylation From Candidate Gene Studies (*n*=18)

| First Author,<br>Year | N | SEP Indicator |  | DNA Methylation |  | Effect<sup>a</sup> |  |  |  |
| --- | --- | --- | --- | --- | --- | --- | --- | --- | --- |
|  |  | Exposure<br>Age<sup>b</sup> | # of<br>Measures | Assessment<br>Age<sup>b</sup> | Targeted<br>Gene(s) | ↑ ↓ | Associated<br>Gene | Associated<br>SEP Domain | Significance<br>Threshold |
| King, 2015 | 489 | Prenatal | 2 | Birth | 9 genes | ↓ | IGF2 | Ed./In. | $p < 0.05$ |
|  |  |  |  |  |  |  | H19 | Ed. |  |
|  |  |  |  |  |  |  | MEG3 | Ed./In. |  |
| King, 2016 | 489 | Prenatal | 1 | Birth | MEG3 | ↑ | MEG3 | Neigh. | $p = 0.002$ |
| Appleton, 2013 | 444 | Prenatal +<br>Birth | 6 | Birth | HSD11B2 | ↓ | HSD11B2 | Comp. | $p < 0.05$ |
|  |  |  |  |  |  |  |  | Ed. |  |
|  |  |  |  |  |  |  |  | Misc. |  |
| Piyasena, 2016 | 50 | Birth | 1 | Birth +<br>Child 2x | IGF2; H19;<br>FKBP5 | ↓ | IGF2<br>FKBP5 | Comp. | $p < 0.05$ |
| Obermann-Borst,<br>2012 | 120 | Child | 1 | Child | IGF2; IGF2R;<br>INSIGF | ↑ | INSIGF | Ed. | $p = 0.021$ |
| Obermann-Borst,<br>2013 | 120 | Child | 1 | Child | LEP | ↑ | LEP | Ed. | $p = 0.008$ |
| Wrigglesworth,<br>2019 | 33 | Child | 1 | Child | BDNF IV | ↑ | BDNF IV | Neigh. | $p = 0.0001$ |
| Huang, 2016 | 613 | Birth | 4 | Adult | 5 genes | ↓ | ABCA1 | Occ. | $p = 0.03$ |
| | | | | | | | HSD11B2 | Ed. | $p = 0.01$ |
| McDade, 2017 | 494 | Child 4x | 1 | Adult | 114 genes | ↑ | GNG2 | Misc. | $p = 0.0093$ |
|  |  |  |  |  |  | ↓ | C1S |  |  |
| Loucks, 2016<sup>c</sup> | 141 | Child | 1 | Adult | 198224 CpGs | ↑ ↓ | 162 CpGs | Comp. | $p < 0.0001$ |
| Needham, 2015 | 1264 | Life<br>course | 3 | Adult | 18 genes | ↑ | 7 genes | Ed. (Child) | $p < 0.05,$<br>$q < 0.2$ |
|  |  |  |  |  |  |  | 6 genes | Ed. (Adult) |  |

|  |  |  |  |  |  |  | 10 genes | Ed. |  |
| --- | --- | --- | --- | --- | --- | --- | --- | --- | --- |
| Smith, 2017 | 1226 | Adult | 2 | Adult | 18 genes | ↑ ↓ | 12 genes | Neigh. | $p \leq 0.05$,<br>$q \leq 0.2$ |
| Stringhini, 2015 | 857 | Life course | 3 | Adult | 17 genes | ↑ ↓ | 2 genes | Occ. (Adult) | $q \leq 5.10 \times 10^{-3}$ |
| | | | | | | ↓ | 6 genes | Occ. | $q \leq 1.49 \times 10^{-3}$ |
| Jones-Mason, 2016<sup>d</sup> | 100 | Adult | 1 | Adult | SLC6A4 | ↑ | SLC6A4 | Comp. | $p = 0.023$ |
| Kogan, 2019 | 309 | Adult | 1 | Adult | OXTR | ↑ | OXTR | Comp. | $p < 0.01$ |
| de Rooij, 2012 | 675 | Adult | 2 | Adult | GR 1-C | ↓ | GR 1-C | Ed. | $p = 0.03$ |
| Simons, 2017 | 100 | Adult 3x | 1 | Adult | OXTR | ↑ | OXTR | Comp. | $p \leq 0.01$ |
| Swift-Scanlan, 2015 | 48 | Adult | 1 | Adult | COMT | ↓ | COMT | Comp. | $p = 0.049$ |

Studies presented in this table are shown in order of DNAm assessment age, then by SEP exposure age followed by alphabetically.
For individual-level study details, see **Table S2**.
Abbreviations: Comp., composite; CpGs, CpG sites; DNAm, DNA methylation; Ed., education; In., income; Occ., occupation; Misc., miscellaneous (i.e., “other” domain); Neigh., neighborhood; SEP, socioeconomic position.
<sup>a</sup>Effects reported at the most stringent significance threshold within the simplest (or unadjusted) model. General direction of effect for association between DNAm and SEP measure reported by arrows, indicating increased or decreased DNAm levels for low SEP.
Associated SEP domain reported with exposure age provided in parenthesis if both child and adult SEP exposures were analyzed. *P*-values reported for significance threshold; *q*-values indicate significance threshold after *p*-values were corrected for multiple testing using the false discovery rate (FDR) method.
<sup>b</sup>SEP exposure age and DNAm assessment age are reported by life course group: prenatal (<0 years), birth (~0 years), child (0-18 years), adult (18+ years). Number of measurements indicated (e.g., 2x) if measured more than once per life course group. “Life course” indicates ages of exposure spanned prenatal, birth or childhood to adulthood.
<sup>c</sup>Loucks et al. 2016 study was included in candidate gene section because study assessed SEP-DNAm associations only in CpG sites that were previously associated with BMI in an EWAS using the same sample.
<sup>d</sup>Reported effect was found when sample was stratified by attachment styles (see Jones-Mason et al. 2016 for more details).

**Table 3.**
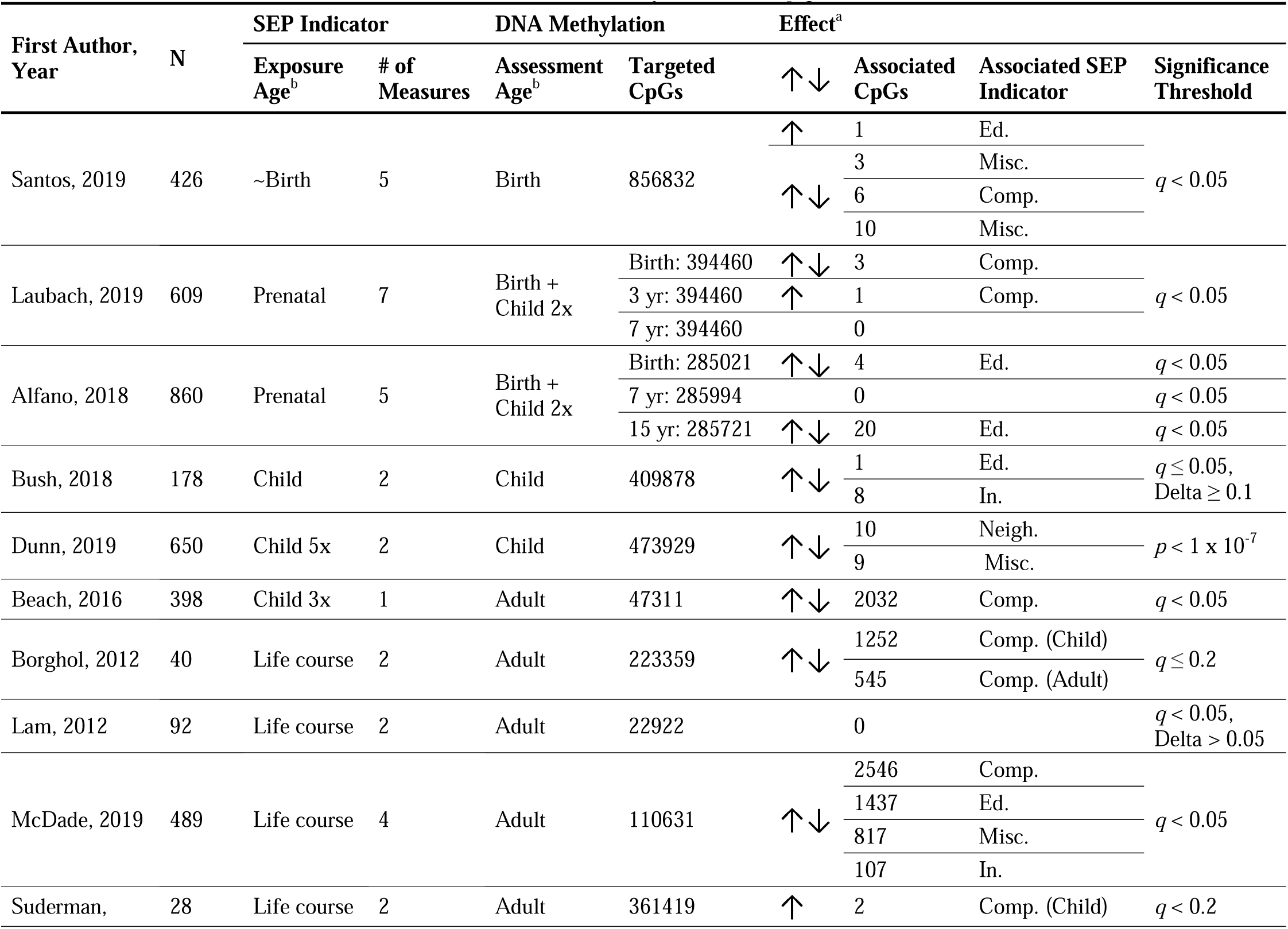
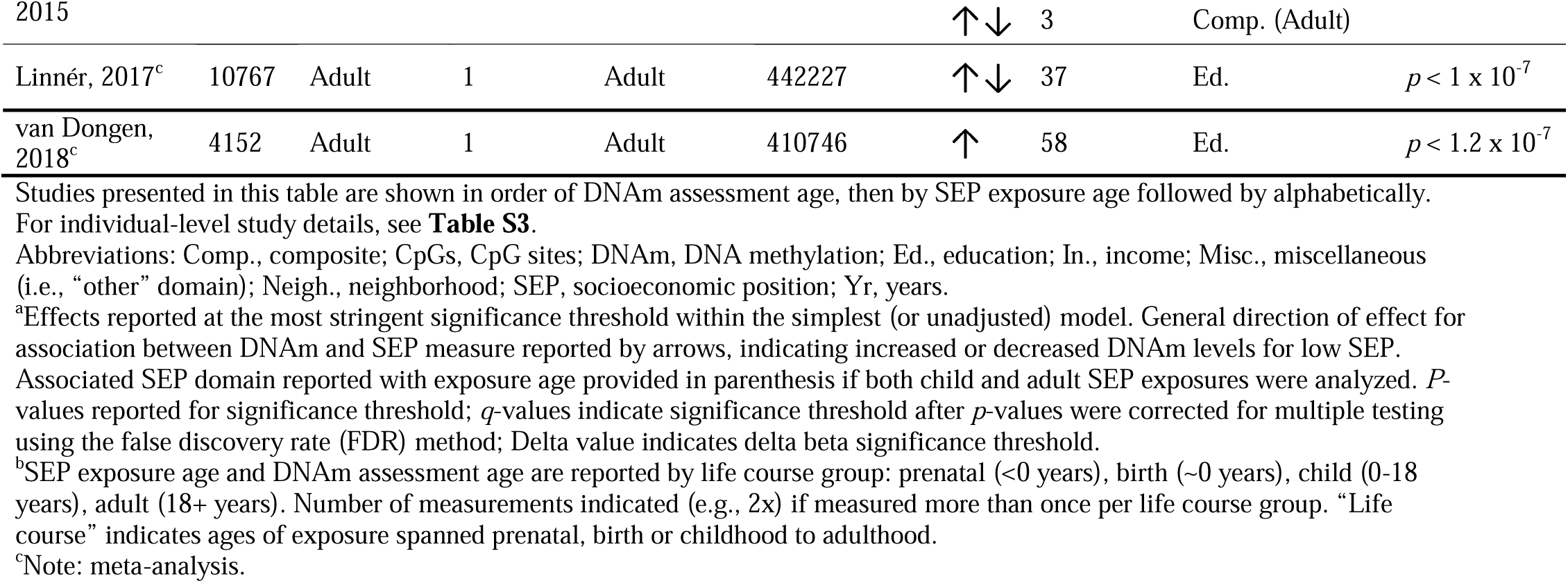
Associations Between Socioeconomic Position and DNA Methylation From Epigenome-Wide Association Studies (*n*=12)

### Overall Study Characteristics

#### Sample features

Nearly all studies (92%; n=34) analyzed samples from cohort studies, wherein participants were observationally followed with data collected either retrospectively or prospectively over a period of time. Specifically, 39 distinct cohorts were sampled in the current review, of which 16 were birth cohorts. Sample sizes varied widely for each study, ranging from 28 to 1264 participants (mean=400). Studies were generally balanced with respect to sex (on average 55% female), although 8 studies included primarily (more than 70%) or entirely female samples and 4 studies included primarily or entirely male samples. Over half of the studies (54%; n=20) sampled participants solely from the United States, while others covered populations from Europe, Canada, Australia, the Philippines, Colombia, and Israel. Most studies focused on multiethnic samples (27%; n=10) or White/majority White samples (24%, n=9), while the remaining included exclusively non-White samples (19%; n=7) or Jewish ethnicity (3%; n=1). The remaining 10 studies (27%) did not directly report race/ethnicity for their sample.

#### Overarching research design

Most studies focused on associations between SEP and DNAm at a single time point, analyzed either cross-sectionally (43%; n=16) or prospectively (43%; n=16); the remaining 14% (n=5) were longitudinal, assessing the same SEP exposure(s) repeatedly across time (**Figure S3**). Of note, two cross-sectional meta-analyses (analyzing cohort-level summary statistics) on the association between adult educational attainment and DNAm were included. Slightly more than half of the studies included (54%; n=20) captured SEP exposure either prenatally, at birth, or during childhood, with another nine (24%) focusing on SEP in adulthood. The remaining eight (22%) studies captured exposure to SEP across the life course (i.e., spanning prenatal, birth, or childhood to adulthood), although half of these studies measured childhood SEP retrospectively in adulthood.

#### SEP exposure features

Across all 37 studies, a total of 96 SEP indicators were individually analyzed, tapping seven different domains: education (n=28 indicators), composite (n=17), occupation (n=12), other (e.g., crowded dwelling, household assets, poverty status; n=13), neighborhood (n=12), income (n=10), and subsidy (i.e., eligible for a form of public assistance; n=4) (**Figure S4**). The number of SEP indicators analyzed across studies ranged from 1 to 7, with a median number of 2. Of the 17 studies analyzing composite measures, 35% (n=6) additionally analyzed each indicator within the composite. Education-related measures were most commonly assessed for all three DNAm approaches (**Figure S4A**). Childhood, including birth, was the most common life-course period examined for SEP exposure (34% of indicators), most often through indices of parent-or household-level education, followed by the prenatal period (32%) and adulthood (27%; **Figure S4B**). Only 6% of indicators captured life-course SEP, spanning early life to adulthood. The majority of SEP indicators were collected objectively through caregiver (52%), self-(30%), or multigenerational reports (< 1%); 11% of indicators were assessed by census-tract data (**Figure S3C**). SEP indicators varied with respect to the measurement scale used to classify individual low to high SEP status for analysis, yet dichotomous measures (47%) were most common (**Figure S4D**).

#### Approach to analyzing DNAm

Candidate gene studies (49%; n=18) were the most common study design, followed by EWAS (32%; n=12) and global DNAm studies (19%; n=7). Most studies assessed DNAm at a single time point in the life course: adulthood (57%; n=21), childhood (19%; n=7), or birth (12%; n=5). Four studies (11%) assessed DNAm repeatedly: at birth and in childhood. Whole blood was the most commonly studied tissue type, used in nearly 30% of studies (n=11) (**Figure S5**). Although most studies targeted one tissue type, five studies (14%) sampled two different tissues to compare between DNAm levels in their analyses.

### The Overall State of the Evidence on the Relationship Between SEP and DNAm

#### Global DNAm studies (n=7)

Within the global group, studies used six different methods for DNAm analysis (**Table S1**). Most global DNAm studies (71%; n=5) reported an association between at least one SEP indicator and global DNAm (**Table 1**). The four studies that measured LINE-1, Alu, or Sat-2 repetitive elements (i.e., a general method for estimating DNAm) reported an association with SEP, whereas only one of the studies measuring DNAm by other global methods reported an association. The significance thresholds and directions of effects were inconsistent between studies.

#### Candidate gene association studies (n=18)

Within the candidate group, studies targeted a variety of gene regions in a total of 274 (unique) candidate genes using four different methods for DNAm analysis; the majority measured DNAm with the MassARRAY EpiTyper (n=8) followed by the 450k array (n=6; **Table S2**). Candidate genes spanned various domains of functional and biological significance, including BMI, stress and inflammation, appetite regulation, fat metabolism, and cardiometabolic processes. All candidate gene studies reported an association between at least one SEP indicator and DNAm for at least one candidate gene, although with inconsistent significance thresholds and direction of these effects across studies (**Table 2**). Two stress-related genes, *OXTR* and *FKBP5*, were the genes most often studied (targeted by three studies each), with all studies reporting increased DNAm for *OXTR* while the direction of DNAm difference for *FKBP5* was mixed across studies.

#### Epigenome-wide association studies (EWAS; n=12)

The majority of EWAS used the 450k array (n=9; **Table S3**). Overall, studies in this category reported 23 different associations between SEP and DNAm, with 8912 total hits passing their most stringent significance thresholds; reporting unique CpGs across studies was impossible due to lack of consistent reporting across studies (**Table 3**). The general direction of DNAm for lower SEP values also varied between studies, with 2685 hits showing increased methylation, 1825 decreased methylation, and the remaining 4402 representing a mix of both (note: DNAm direction for the remaining hits were unreported at the individual-level for simple models).

### Does the Timing and/or Duration of SEP Influence the Association Between SEP and DNAm?

The majority of studies covered in this review examined a single life-course period of SEP exposure in relation to DNAm. However, nine studies (two global DNAm (44, 45), two candidate gene (24, 25), and five EWAS (22, 23, 66, 69, 71)) investigated how the timing and/or duration of SEP exposure impacts the relationship between SEP and DNAm. These studies found evidence that the timing and duration of SEP may influence the association between SEP and DNAm (**Table 4**), although the magnitude and direction of these timing effects were inconsistent across studies.

**Table 4.** Overview of Findings From Studies (*n*= 9) Investigating the Timing and/or Duration of SEP and DNA Methylation

| First Author, Year | DNAm Approach | SEP Exposure Age<sup>a</sup> | Reported Age at DNAm<sup>b</sup> | Key Findings |
| --- | --- | --- | --- | --- |
| Sensitive period of child SEP on child or adult DNAm |  |  |  |  |
| Dunn, 2019 | EWAS | Child (prospectively measured 5x) | 7 | Neighborhood disadvantage and financial stress associated with 10 and 9 CpGs ($p < 1 \times 10^{-7}$) respectively, with 8 and 2 of these differentially methylated sites relating to the developmental timing of SEP exposure in very early childhood (birth to age 2). |
| Lam, 2012 | EWAS | Child (retrospectively measured) + Adult | Mdn=33.04 | Low early life SEP associated with 3 CpGs ($q < 0.25$), while current (adult) SEP did not. |
| Borghol, 2012 | EWAS | Child (prospectively measured) + Adult | 42 + 45 | Child SEP associated with 1252 gene promoters, while adult SEP associated with 545 ($q \leq 0.2$). Child and adult SEP-associated promoter sets overlapped at 63 promoters. |
| Sensitive period of adult SEP on adult DNAm |  |  |  |  |
| Stringhini, 2015 | Candidate Gene | Child (retrospectively measured) + Adult | ~ 45–63 | Current adult SEP associated with 2 genes ($q < 0.001$), while child SEP did not have any associations. |
| Subramanyam, 2013 | Global DNAm | Child (retrospectively measured) + Adult | M=61 | One SD higher adult wealth associated, on average, with 0.09% higher Alu ($p < 0.01$) and 0.15% lower LINE-1 DNAm ($p < 0.01$), while adult income, adult education, and child SEP did not associate with LINE-1 or Alu DNAm. |
| Sensitive periods of both child and adult SEP on adult DNAm |  |  |  |  |
| Tehranifar, 2013 | Global DNAm | Child (prospectively measured) + Adult | M=43 | Lower maternal education and lower family income associated with higher mean levels of Sat-2 DNAm ($p < 0.05$ and $p < 0.10$, respectively). Single-parent family structure compared to two-parent structure associated with higher Alu methylation ($p < 0.05$). Adult income associated with increased LINE-1 methylation ($p < 0.05$). |
| Suderman, 2015 | EWAS | Child (prospectively measured) + Adult | 45 | Child SEP associated with 2 CpGs, while adult SEP associated with 3 CpGs ($q < 0.2$) in whole blood or LCLs. None of the SEP-associated CpGs were shared between child and adult SEP. |
| Needham, 2015 | Candidate Gene | Child (retrospectively measured) + Adult | M=69.55 | Low child SEP associated with DNAm in 3 stress- and 2 inflammation-related genes, whereas adult SEP primarily associated with DNAm in inflammation-related genes (5 inflammation- and 1 stress-related gene; all $p < 0.05$; $q < 0.20$). SEP-DNAm associations between child and adult SEP overlapped at 3 of 9 total candidate genes. |
| Effects of life-course SEP trajectories on adult DNAm |  |  |  |  |
| McDade, 2019 | EWAS | Child + Adult (prospectively measured 4x) | M=20.9 | Persistently low SEP from infancy to young adulthood associated with 2546 CpGs ($q < 0.05$), compared to persistently high SEP. One CpG associated with DNAm for upward SEP mobility group and no sites associated with the downward mobility group ($q < 0.05$). |
| Stringhini, 2015 | Candidate Gene | Child (retrospectively measured) + Adult | ~ 45–63 | Persistently low SEP from childhood to adulthood associated with 6 genes, compared to persistently high SEP, while downward SEP and upward SEP mobility groups associated with 4 and 1 genes, respectively ($q < 0.001$). |
| Needham, 2015 | Candidate Gene | Child (retrospectively measured) + Adult | M=69.55 | Persistently low SEP from child to adulthood associated with DNAm in 5 genes, upward SEP mobility associated with 1 gene, and downward mobility associated with 2 genes in comparison to persistently high SEP ($p < 0.05$; $q < 0.20$). |

Studies presented in this table are categorized by general findings in support of sensitive periods of child SEP, adult SEP, or both and DNAm, and by findings from SEP trajectory studies. Within these categories, studies are shown in descending order of DNAm assessment age.
Abbreviations: CpGs, CpG site; DNAm, DNA methylation; LCLs, lymphoblastoid cell lines. SEP, socioeconomic position; SD, standard deviation.
<sup>a</sup>SEP exposure age column reports the life course period of exposure captured (prenatal, child, adult) and notes whether measure was prospectively or retrospectively assessed. Number of measurements indicated (e.g., 2x) if same SEP indicator was measured more than once.
<sup>b</sup>Reported age at DNAm age column reports the age of DNAm assessment in years (as reported by individual studies).

With respect to findings on the relative importance of exposure timing, two (22, 66) of seven studies comparing child and adult SEP found stronger support for sensitive periods of child SEP with adult DNAm changes as compared to adult SEP. By contrast, two (25, 44) of these studies found stronger support for adult SEP associations with adult DNAm changes compared to child SEP, noting that the lack of associations for child SEP may be due to measurement limitations (e.g., retrospectively assessed, limited SEP variability). The remaining three studies (24, 45, 71) found support for both child and adult SEP associations with adult DNAm changes, observing diverse methylation changes between child and adult indicators. Only one study (23) analyzed repeated indicators of low SEP at different childhood periods in relation to childhood DNAm patterns; findings from that study suggested sensitive-period effects, such that low SEP in very early childhood (before age 3) was associated with 10 of 19 differentially methylated CpGs at age 7 (*p* < 1 × 10^−7^).

Three studies in this group (24, 25, 69) also captured effects of life-course SEP trajectories on adult DNAm, such as moving from low child to high adult SEP (**Table 4**). These studies consistently found DNAm differences between persistently low SEP (both low child and low adult SEP) compared to persistently high SEP groups, with fewer or no DNAm differences observed for either upward (moving from low child SEP to high adult SEP) or downward mobility groups.

Taken together, these findings provide evidence for an effect of SEP timing and duration on DNAm; this relationship may be unique for SEP indicators measured in childhood and persistent exposure to low SEP across the life course. Furthermore, the unique DNAm patterns observed for child versus adult SEP suggest that SEP exposure may potentially operate through different underlying biological pathways across development.

### Do Different SEP Indicators Associate Differently with DNAm Profiles?

We addressed our second research question in two parts with compiled summary statistics of the nine EWAS studies included in the current review that used the 450k array (**Supplemental Materials**). In part one, we found 113 unique CpGs (FDR < 0.05) in two or more studies, with five CpGs appearing between three different studies (**Table S4**). **Figure 1** presents the pattern of overlap in the 113 CpGs across four different SEP domains, with five CpGs associated with all domains. There were 264 total associations between SEP and DNAm across domains; education had the highest number of associations (n=95), of which 54 (57%) were unique loci.

**Figure 1.**
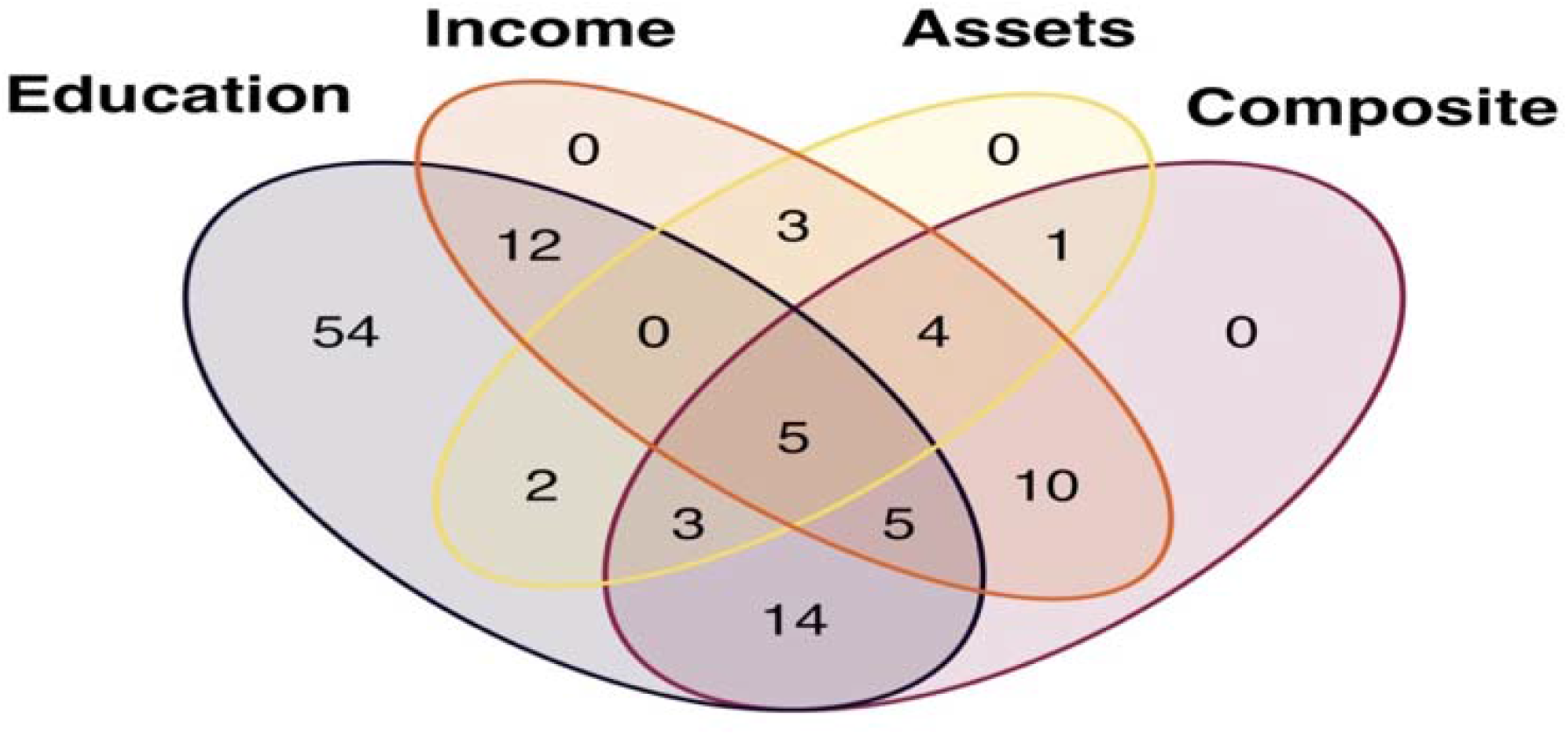
Venn diagram illustrating the overlap of unique, top CpG sites (CpGs) across socioeconomic position (SEP) domains Pattern of overlap in 113 significant SEP-associated CpGs (FDR<0.05) appearing in more than one study across four SEP domains: education, income, assets (household), and composite. As shown for income, there were no unique associations found among the 39 CpGs predicted by income, with 12 CpGs overlapping with education, 10 with composite, 3 with assets, and the remaining 14 overlapping with two or more domains. CpG-level data was compiled from summary statistics of nine epigenome-wide association studies utilizing the Illumina Human Methylation 450k array. For more information on how these summary statistics were derived, see **Supplemental Materials**. For a list of 113 associated CpGs IDs, see **Table S5**.

In part two, we filtered the summary statistics to the CpGs analyzed across all studies at an FDR < 0.05. Few CpGs were shared between the four SEP domains (**Figure 2**). A total of 3670 CpG associations survived FDR adjustment and more than half (n=2002; 55%) were unique to a single SEP domain. Composite measures were linked to the highest total number of significant CpGs (1389), 652 of which (47%) were unique. Education was associated with the second highest total number of CpGs (1114), 686 of which (62%) were unique. A total of 623 associated CpGs were reported for income and 544 for assets, of which 548 (88%) and 116 (21%) were unique, respectively. Overall, these results suggest that different SEP indicators, particularly education and income, may represent distinct aspects of the socioeconomic environment and thereby present unique relationships with DNAm.

**Figure 2.**
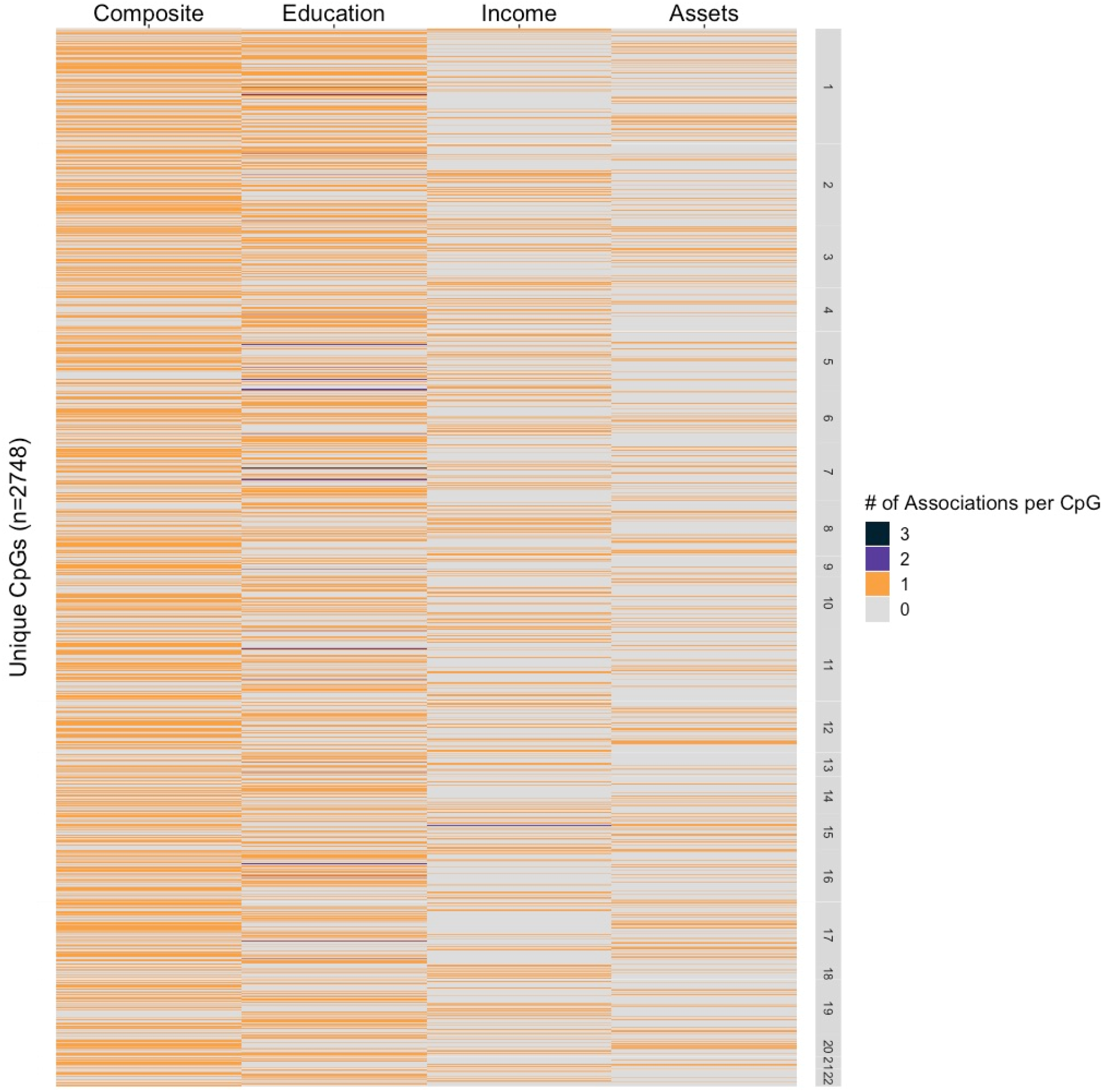
Heat map demonstrating the overlap of shared, top CpG sites (CpGs) across socioeconomic position (SEP) domains This figure presents a heat map of the overlap in CpGs identified as signficantly associated (FDR<0.05) with SEP indicators categorized by four domains: composite, education, income, and assets (household). To produce this heat map, we first collected individual CpG-level summary statistics with corresponding SEP indicator(s) and *p*-values for 8 epigneome-wide association studies utilizing the Illumina Human Methylation 450k array, adjusting for FDR within each study. We then subset the full CpG list to: (a) only the CpGs analyzed across all studies, and (b) CpGs that survived FDR<0.05 adjustment. Through these two filters we arrived at 2748 unique SEP-associated CpGs between 6 studies. Next, we assigned every CpG a value based on the number of significant associations (FDR < 0.05) per SEP domain. Colors indicate the number of associations per CpG per SEP domain, ranging from 0 to 3. For each SEP domain, a CpG received a value of 0 if it did not survive FDR adjustment or was not analyzed in that domain, a value of 1 if it was significantly associated in just one study, a value of 2 if it was significantly associated between two studies (within the same domain), and so on. To interpret these results, we plotted individual CpGs along the y-axis by chromosomal position and grouped SEP indicators into four primary domains along the x-axis, in columns. The heatmap shows no apparent pattern in CpGs by chromosome. For shared associations between studies, 59 CpGs appeared in two different studies and 5 CpGs appeared in three different studies, with the remaining 2684 CpGs appearing in only one study. For associations shared between more than one study in each column, 36 CpGs associated with education between two studies and 3 CpGs associated with education between three studies. In the income domain, one CpG associated between two different studies. No CpGs were shared between studies for composite and assets. Composite measures associated with the highest number of CpGs (n=1389), followed by education (n=1156), income (n=624), and assets (n=544). There appears to be little overlap in CpGs between domains, with 88% of CpGs in the income domain having unique signal, followed by 62% for education, 47% for composite, and 21% for assets. The comparison of overlap in CpGs between SEP domains is limited by the variation in the number and type of indicators analyzed by each study, which varied from 1-4 indicators within each study (see **Table S4** for more details on summary statistics).

## Discussion

Three main findings emerged from this review. First, indicators of child and adult SEP shared little overlap in adult DNAm profiles, suggesting that SEP can become biologically embedded through distinct and potentially time-dependent pathways across development. Such findings are consistent with prior life-course research showing that risks for adverse health outcomes differentially arise from child and adult SEP (73). For example, behavioral and health risk factors (e.g., cigarette smoking, low exercise levels) are more strongly predicted by adult SEP, while physiological risk factors (e.g., BMI, cardiovascular disease) are more strongly associated with child SEP (74). However, less than 25% of studies included here compared the associations between child and adult SEP and adult DNAm changes. In fact, nearly half of studies captured child SEP retrospectively during adulthood, with all measuring DNAm cross-sectionally in adulthood. Building from these findings, longitudinal data from birth cohorts is needed to analyze prospective measures of SEP and repeated measures of DNAm to determine whether differences in DNAm appear early in life, later in adulthood, or change over the life course.

Second, suggestive evidence emerged for SEP timing and duration effects on DNAm, consistent with life-course theories on mobility (75, 76), sensitive periods (77, 78), and accumulation of risk effects (79, 80). Three trajectory studies evaluating mobility, the most commonly studied life-course theory in the current review, found differences in DNAm profiles between groups exposed to persistently low compared to persistently high SEP from childhood to adulthood. These findings are consistent with prior studies showing cumulative effects of socioeconomic disadvantage on poor health outcomes into adulthood (73, 81, 82). Of importance, findings from these trajectory studies also showed that upward/downward SEP mobility groups had fewer DNAm differences compared to persistently low SEP individuals, suggesting that early-life DNAm patterns may not be fixed in development, but rather that SEP effects might be alleviated through upward mobility in later life (83, 84). However, these trajectory studies were few in number and the SEP trajectories analyzed were limited to two timepoints. Only one study tested a sensitive period hypothesis at multiple stages in childhood, representing the first time a sensitive period model was applied in an epigenome-wide context. There, SEP reported in the first three years of life was the strongest predictor of DNAm differences at age 7, as compared to later in development (23). These findings highlight that very early life may be a particular sensitive period for the ill effects of socioeconomic disadvantage (85, 86).

Third, we found little overlap in DNAm patterns across SEP domains, suggesting that different SEP indicators likely represent different aspects of the socioeconomic environment and thus, may leave distinct biological signatures. Past reviews have examined the relationship between multiple SEP indicators and health outcomes, noting that SEP indicators are independent from each other, and that measures such as education and income are not interchangeable (87, 88). Yet, education and income were most commonly investigated across studies, leaving other key SEP indicators such as neighborhood-level SEP relatively absent in the broader epigenetic literature, despite their effects on numerous health outcomes (89). This gap highlights the need for future epigenetic studies to more evenly capture SEP operating at different socioecological levels, in order to elucidate the potentially different downstream health effects of various SEP exposures (90). Moreover, there was little consensus in how studies measured SEP, with over 40 different definitions examined. In particular, few consistencies were observed across SEP indicators that were used to capture between-person variability within similar population demographic groups. Without greater consensus on best practices in defining SEP (3), comparisons between DNAm outcomes will remain challenging to interpret, and potential differences between indicators at the same level (individual-, household-, and neighborhood) will be difficult to discern.

Given these findings, what are some promising directions for future research? There is clearly a need for more longitudinal research with repeated measures of both SEP and DNAm, which will allow researchers to test targeted life-course theories and explore causal pathways about how SEP becomes biologically embedded (91). It is also crucial to more precisely conceptualize and measure SEP, which can be achieved through 1) thoughtfully selecting SEP variables that better represent levels of SEP in a given population (e.g., indicators of wealth should be prioritized in elderly populations (92)), 2) explicitly defining SEP measures in terms of the components captured, and 3) analyzing a comprehensive set of SEP indicators (i.e., across different domains and levels), including the individual components of an aggregated composite measure. With these innovations described above, researchers will gain a clearer picture of not only *whether*, but also *when* and *how* different dimensions of the socioeconomic environment influence DNAm and possibly other biological processes.

## Conclusion

As socioeconomic inequality continues to grow on a global scale (93), the importance of understanding the health consequences of socioeconomic position increases worldwide. SEP is a fundamental social determinant, influencing nearly all aspects of the environment that contribute to overall health, and must be considered in epigenetic studies of social and behavioral traits (94), whether as a control or independent variable. To better understand how the socioeconomic environment interacts with the epigenome and other biological processes to contribute to health disparities, researchers must consider the implications and limitations of evidence due to the diversity of SEP measures, while also applying rigorous design and analytic approaches that allow for the investigation of SEP timing, duration, and type. With these efforts, we can tackle the complexities of how SEP becomes biologically embedded and help guide future intervention and prevention strategies to effectively reduce SEP-related health disparities across diverse populations.

## Data Availability

N/A

## Acknowledgements

The Russell Sage Foundation (Grant ref: Trustee Grants, Integrating Biology and Social Science G-6692) provided core support for this research. This work was also supported by the National Institute of Mental Health of the National Institutes of Health (E.C.D., Award Number R01MH113930). This publication is the work of the authors, each of whom serve as guarantors for the contents of this paper.

