## Supplemental Materials for "Associations between indicators of socioeconomic position and DNA methylation: A systematic review"

Study Identification

*Full Search Terms*

We systematically searched for articles published from inception through September 18, 2019 on PubMed and PsycINFO by using a combination of index/MeSH terms (i.e., keywords) and individual terms located in the title or abstract that were related to socioeconomic position (SEP) and DNA methylation (DNAm). To capture the multifaceted nature of SEP, we included terms for the following domains: a) education b) occupation (including employment), c) income (including financial difficulties), and d) neighborhood characteristics. No language, publication date/status, or age restrictions were imposed. The following were the final search terms used to search for articles in PubMed: *(“Socioeconomic Factors”[Mesh] OR “Income”[Mesh:NoExp] OR Income[tiab] OR Income distribution[tiab] OR Living standard*[tiab] OR Standard of living[tiab] OR Socioeconomic ­­status*[tiab] OR Socioeconomic factor*[tiab] OR Occupation[tiab] OR Vocation[tiab] OR Social class*[tiab] OR Poverty area*[tiab] OR Neighborhood disadvantage[tiab] OR Neighborhood characteristics[tiab] OR adversity[tiab]) AND (“DNA Methylation”[Mesh] OR “Epigenesis, Genetic”[Mesh] OR “Epigenomics”[Mesh] OR DNA Methylation*[tiab] OR Epigenet*[tiab] OR Epigenetic Process*[tiab] OR Genetic epigenesis[tiab]).* Terms followed by “[Mesh]” were index terms used to search by subject content of indexed journal articles. Individual terms followed by “[tiab]” searched for articles that included the term in the abstract or the title; * denotes truncation.

In PsycINFO database, the following were the final search terms used to search for articles in PsycINFO: *((DE "Socioeconomic Status" OR DE "Family Socioeconomic Level" OR DE "Income Level" DE "Social Class"OR DE "Parental Occupation" OR DE "Occupational Status" OR DE "Occupational Mobility" OR DE "Occupations" OR DE "Occupational Success"OR DE "Poverty" OR DE "Social Deprivation" OR DE “Disadvantaged” OR DE “Poverty” OR DE “Income (Economic)” OR DE “Income Level” OR DE “Financial Strain” OR DE "Educational Attainment Level") OR TI (“social class” OR “socioeconomic*” OR “income” OR “occupation” OR “education”) OR AB (“social class” OR “socioeconomic*” OR “income” OR “occupation” OR “education”) OR SU “Socioeconomic Status”)) AND (DE “Epigenetics” OR TI (“DNA Methylation*” OR “epigenetics” OR “methylation”) OR SU “Epigenetics” OR AB (“DNA Methylation*” OR “epigenetics” OR “methylation”)).* Terms following “DE” and “SU” were index terms used to search by subject content of indexed journal articles and by subject heading; individual terms following “TI” and “AB” searched for articles that included the term in the abstract or the title; * denotes truncation.

Study Selection

*Inclusion and Exclusion Criteria*

The search and selection procedure is shown in **Figure S1.** The PubMed and PsycINFO search returned 478 results. Titles and abstracts were reviewed using the following inclusion criterion: human empirical studies that examined an independent association between at least one SEP exposure measure and DNAm as an outcome. Most articles (366) were excluded because they did not include an SEP measure as an exposure and/or did not include DNAm as an outcome. Thirteen articles were removed because they measured DNAm with an epigenetic biomarker of age to estimate “epigenetic age,” or estimates of biological age based on DNAm. Another 10 were removed because they did not include a healthy control group or their sample was homogenous for SEP status (e.g., entirely low income). Four animal studies were removed. Three studies were excluded because they combined SEP and non-SEP (e.g., childhood abuse or parenting stress) measures into one aggregated composite measure. Fifty-five were reviews, overviews, or commentaries and were also excluded. We identified six additional studies by reviewing reference lists of 29 eligible publications and also added two known publications to our review. In final, 37 studies were included in this systematic review.

Data Extraction

*SEP Exposure Features*

The following domains were used to categorize each measure indicating a component of the SEP construct: education, occupation, income, neighborhood, subsidy, composite, and other. All measures relating to *education* (e.g., highest level of education, number of years of schooling, highest degree, etc.) were included in the education domain. Measures related to occupation or employment (e.g., highest level of household occupation, unemployment) were included in the occupation domain. Measures related to *income* (e.g., weekly or annual income) were included in the income domain. Measures of the *neighborhood* domain captured physical, structural, and functional aspects of an individual’s neighborhood through either self-reported questionnaires or by geography/zip code via census-tract data (1). The *subsidy* domain included measures indicating whether individuals were eligible or on a form of public assistance, such as food or housing subsidies. We also created a domain for *composite* measures*,* which included studies that used measures that were either: a) a cumulative score derived by aggregating multiple scores from indicators that spanned more than one domain of SEP (summing, averaging, standardizing, etc.), or b) from pre-existing scales or indexes measuring multiple domains of SEP (e.g., Hollingshead Four Factor Index of Socioeconomic Status). Finally, the *other* domain included miscellaneous indicators of SEP, such as poverty status, crowded dwelling, and difficulty affording household assets.

Ages of SEP exposure were grouped by life-course period: prenatal (<0), birth (0), child (0-18 years), adult (18+ years). Studies that captured SEP spanning early life to adulthood (i.e., prenatal, birth, or child to adult) were classified as “life course” for exposure age. Method of collection was grouped into five categories: caregiver report, self-report, multigenerational (i.e., both caregiver and self-report), census-tract (i.e., taken from government official statistical subdivisions of a county of geographical equivalent), and cohort-level summary statistics. For longitudinal and prospective studies, SEP exposures were also reported as prospective or retrospective for method of collection. Scale of measurement defined how each measure was statistically encoded for analysis, grouped by continuous (continuous numerical scale), dichotomous (binary), categorical (unranked), and ordinal (ranked). For more details on individual study SEP indicators and definitions, see **Tables S1-S3**.

Summary Statistics Analyses and Supplementary Results

We addressed our second research question (do different SEP indicators show differential DNAm profiles?) in two parts with compiled summary statistics of the nine epigenome-wide studies (EWAS) included in the current review that used the Illumina Human Methylation 450k array method (2-10). We adjusted *p*-values using a 5% false-discovery rate (FDR) (11) within each study by each SEP indicator analyzed and annotated CpG sites (CpGs) to the nearest gene (located in the gene body or within 300 kb of a transcription start site, TSS) and chromosome position using the *FDb.InfiniumMethylation.hg19* package in R/Bioconductor. Additional descriptives of the summary statistics by individual study level, including sample size, age at assessments, tissue type(s), and covariates are included in **Table S4**. In part one, we compared top CpGs (FDR < 0.05) between the nine studies. Of note, one of these nine studies (2) was a mediation analysis and only the top CpGs included in the mediator were available for analysis. Before FDR adjustment, there were a total of 482674 unique CpGs spanning seven domains: composite, education, household assets, income, neighborhood, occupation, and other. A total of 7652 unique CpGs survived adjustment for FDR < 0.05 across eight studies and four SEP domains (i.e., composite, education, income, household assets). We found 113 significant CpGs appearing in more than one study, with five appearing between three studies and the remaining between two studies (see **Table S5** for list of individual CpG IDs and annotated genes).

These 113 CpGs had 264 associations across four domains of SEP. The majority of associations were found for education (95 total), followed by composite (42), income (39), and assets (18; **Figure 1**). Almost half (n=53; 47%) of CpGs appeared solely between the two meta-analyses on educational attainment (6, 9), which shared partially overlapping samples. Eight of these 53 sites were previously reported as shared between these two meta-analyses, for adjusted models additionally controlling for smoking (9). In total, CpGs annotated to 73 unique genes, with *AHRR* annotating to the most unique number of CpGs (n=9), followed by *CDK6*, *MYO1G*, and *PRSS23* annotating to four total CpGs each.

In part two, we performed a between-study comparison of all analyzed CpGs predicted by SEP at FDR < 0.05 and assessed the extent of overlap in top CpGs between SEP domains. We excluded the mediation analysis of Beach et al. (2016) and filtered the compiled summary statistics to (a) only CpGs analyzed across all eight studies, and (b) CpGs that survived FDR < 0.05 adjustment. Through these two filters we identified 2748 unique CpGs between six studies (3, 5-7, 9, 10), spanning four SEP domains (i.e., composite, education, income, and household assets). Of note, two of the six studies included in this analysis analyzed composite measures based on other indicators of education, income, and assets (only for McDade et al. 2019) included in **Figure 2**, which may explain the lack of unique signal found for composite and assets.

**Supplementary Figures**

Figure S1. Search and selection procedure

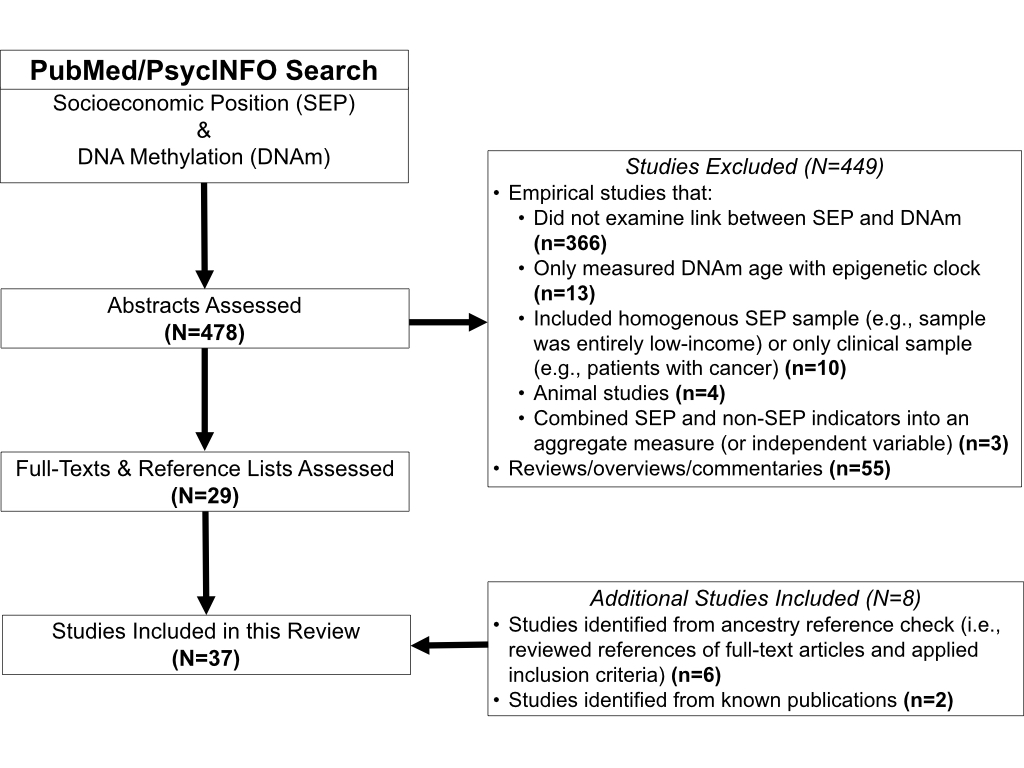

The full search and selection procedure of published studies from inception through September 2019 for a systematic review of the association between socioeconomic position (SEP) and DNA methylation (DNAm). A PubMed and PsycINFO search returned 478 articles. Abstracts were assessed and most articles (366) were excluded because they did not include an SEP measure as an exposure and/or did not include DNAm as an outcome. Thirteen articles measured DNAm with an epigenetic biomarker of age to estimate “epigenetic age,” or estimates of biological age based on DNAm, and were also removed. Another 10 were removed because they did not include a healthy control group or their sample was homogenous for SEP level (e.g., entirely low income). Four animal studies were removed. Three studies were excluded because they combined SEP and non-SEP (e.g., childhood abuse) measures into one aggregated composite measure. Fifty-five were reviews, overviews, or commentaries and were also excluded. We identified six additional studies by reviewing reference lists of 29 eligible publications and also added two known publications to the review. In final, 37 studies were included in this systematic review.

Figure S2. Trends in publications over time

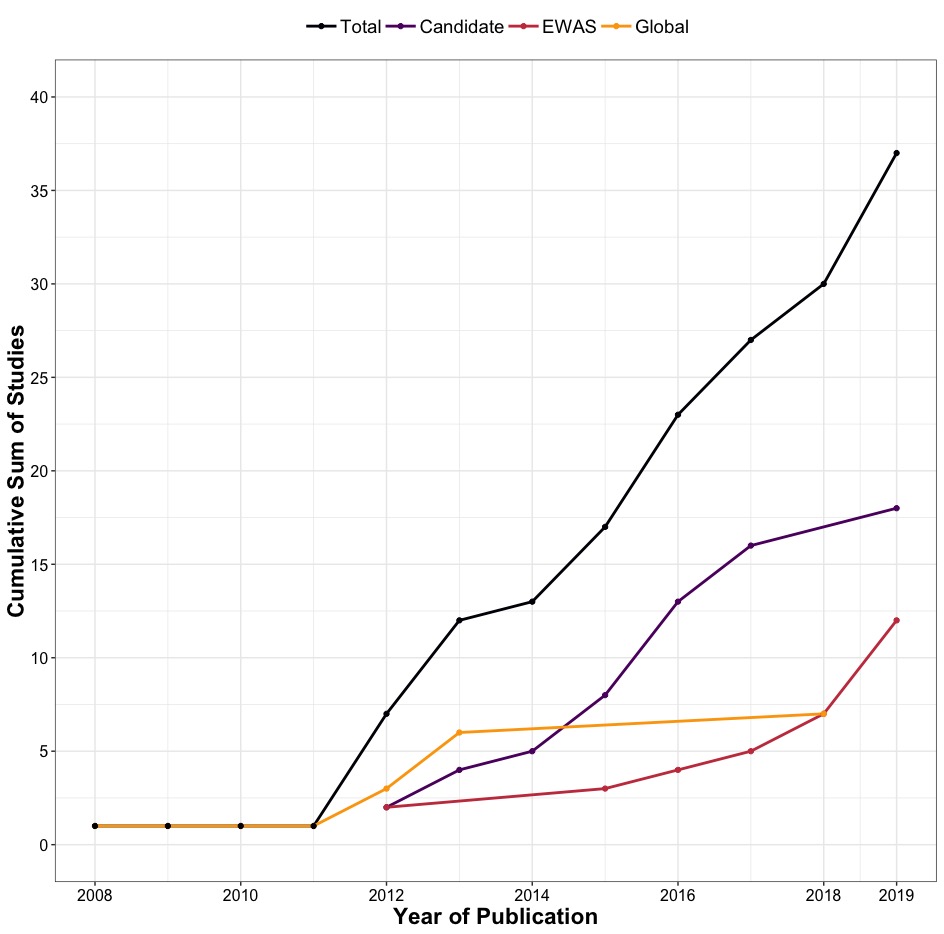

This figure presents the cumulative number of 37 total published studies by year and approach to analyzing DNA methylation (DNAm). We conducted an exhaustive literature search from inception until 09/18/2019 using electronic databases PubMed and PsycINFO. This graph illustrates the 37 papers included in the review, beginning with articles published on or after 01/01/2008, the date of the first included published paper identified by our search.

Total, total number of publications for all types of DNAm approaches; Candidate, candidate-gene association studies; EWAS, epigenome-wide association studies; Global, global DNAm studies.

Figure S3. Assessment of life-course stages

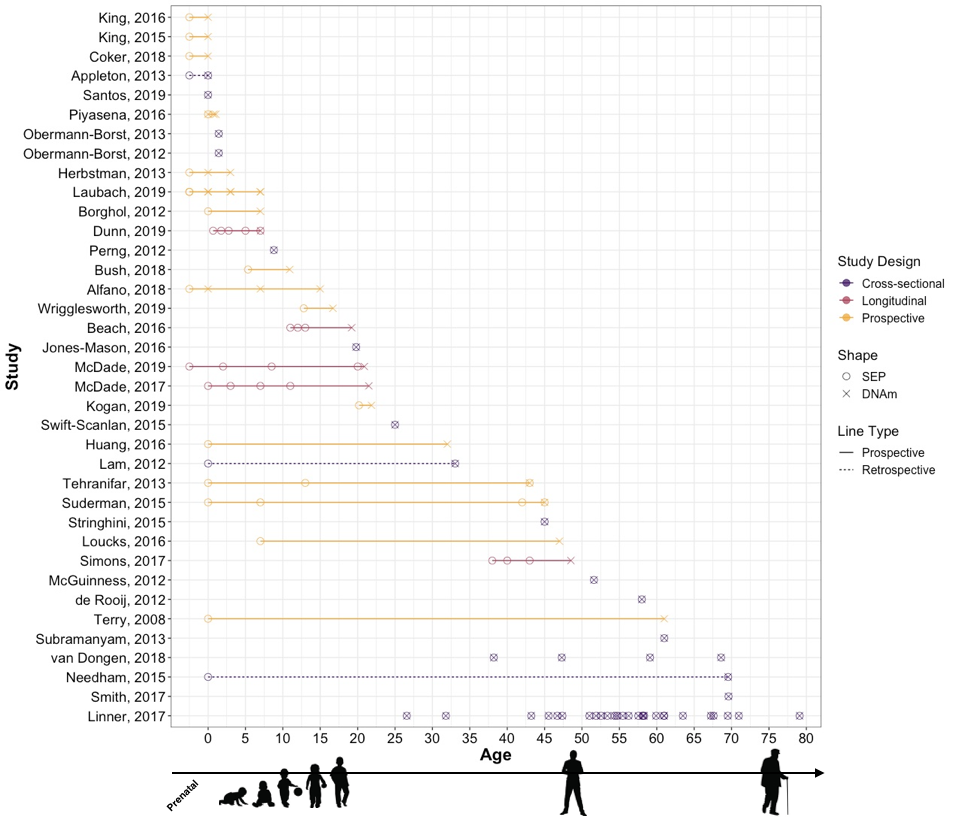

The stages in the life course captured by socioeconomic position (SEP) exposure age and age at DNA methylation (DNAm) assessment are plotted by study design (cross-sectional, prospective, longitudinal) across all 37 studies included in review. Life-course stages include prenatal (<0), birth (0), child (0-18 years), adult (18+ years).

Cross-sectional, study captured SEP exposure(s) and assessed DNA methylation at the same time in the life course; Prospective, study prospectively assessed SEP exposure(s) no more than once over the life course; Longitudinal, study prospectively assessed the same SEP exposure(s) at least twice over the life course.

Solid lines indicate SEP was prospectively assessed, while dotted lines indicate SEP was retrospectively captured.

Note: Linnér, 2017 and van Dongen, 2019 were meta-analyses.

Figure S4. Trends in socioeconomic position (SEP) indicators

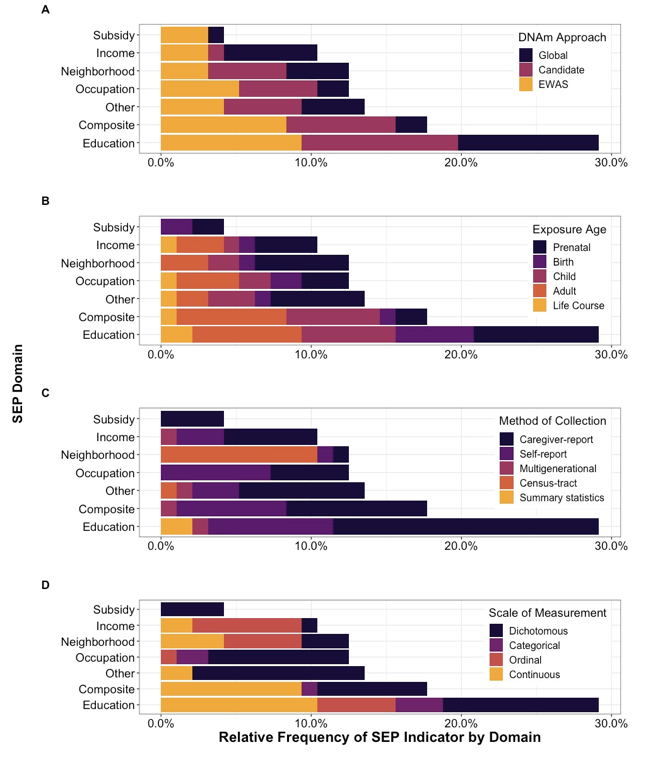

The total number of SEP indicators included in review, categorized by SEP domain and presented by: (A) DNA methylation (DNAm) approach, (B) exposure age, (C) method of collection, and (D) scale of measurement. Relative frequencies are reported, which refer to the percent (%) of SEP indicators by domain over the total number of indicators (n=96) included in the 37 studies examined. For example, measures defined as occupational prestige, employment status, or job title were classified under the *occupation* domain. *Composite* domain included cumulative measure of aggregated SEP measures from multiple SEP domains (e.g., a composite score summarizing occupation, income, and education measures). Measures such as household assets, crowded dwelling, and poverty status were grouped under the *other* domain. For more details on SEP indicators at the individual study level, see **Tables S1-S3.**

(A) Approach to analyzing DNAm categorized into three groups: global DNAm, candidate gene, and EWAS (epigenome-wide association study).

(B) SEP exposure age reported by life-course group: prenatal (<0 years), birth (~0 years), child (0-18 years), adult (18+ years), and life course (ages of exposure captured spanned early life to adulthood).

(C) Method of collection used to collect information on SEP indicator categorized by: caregiver report, self-report, multigenerational (both parent and self-reports), census-tract (taken from government official statistical subdivisions of a county of geographical equivalent), and summary statistics.

(D) Scale of measurement used to code SEP indicator for analysis; scales include dichotomous (binary), categorical (unranked), ordinal (ranked), and continuous (numerical).

Figure S5. Trends in tissue type

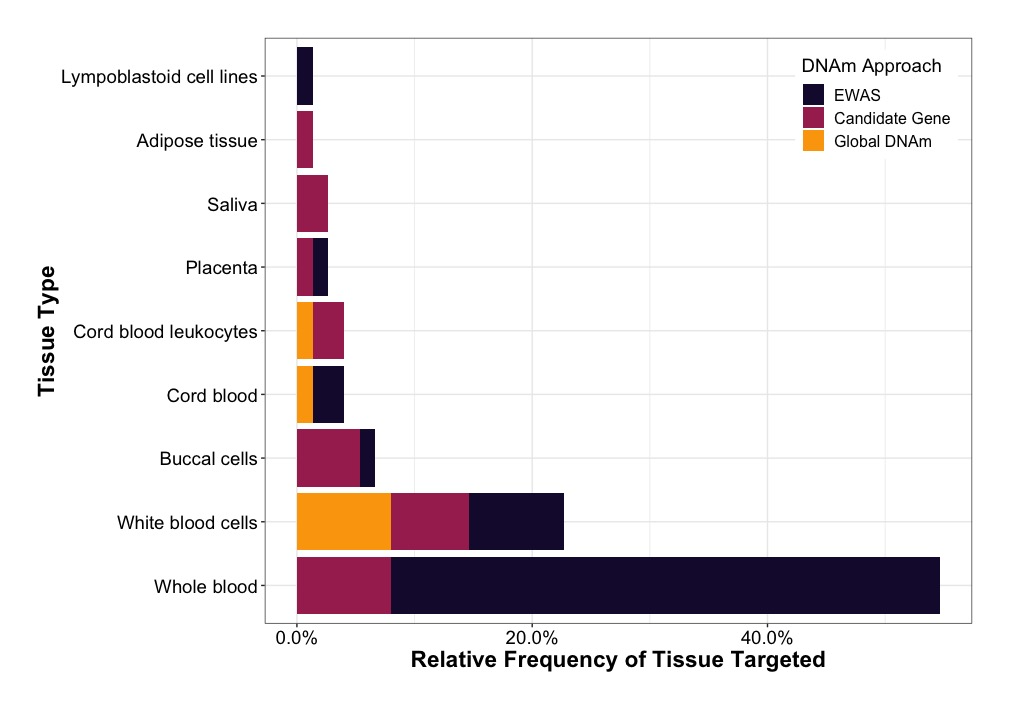

The number of tissue types targeted across 37 studies included in the review, separated by DNA methylation (DNAm) approach: EWAS (epigenome-wide association studies), candidate gene, and global DNAm. Relative frequencies are reported, which refer to the percent (%) out of the 37 studies examined. The most commonly assessed tissue type was whole blood (55%), which was targeted by only EWAS and candidate gene studies, followed by white blood cells (23%), which was evenly targeted between the three DNAm approaches. The remaining tissue types were targeted by less than 1% of studies. Five studies targeted two different tissue types. Tissue types included by each study are presented in **Tables S1-S3.**
